# Prevalence and determinants of mental well-being and satisfaction with life among university students amidst COVID-19 pandemic

**DOI:** 10.1101/2022.05.18.22275203

**Authors:** Md. Safaet Hossain Sujan, Atefehsadat Haghighathoseini, Rafia Tasnim, Rezaul Karim Ripon, Sayem Ahmed Ripon, Mohammad Mohiuddin Hasan, Muhammad Ramiz Uddin, Most. Zannatul Ferdous

## Abstract

**Background:** The COVID-19 pandemic has caused a slew of mental illnesses due to a lack of cures and vaccinations, as well as concerns about students’ well-being and satisfaction with life, resulting in psychological symptoms and dissatisfaction with their lives. As students are highly susceptible to mental health issues, researchers discovered that perceived SWL and MWB decreased. The present study investigated the prevalence and determinants of mental well-being and satisfaction with life among university students in Bangladesh.

**Methods:** An e-survey based cross-sectional study was carried out from February to April 2021 among 660 students. A purposive sampling technique was utilized in the study. Self-reported mental well-being and satisfaction with life psychological tools were also used. The e-questionnaire survey was conducted with informed consent and questions were related to socio-demographics, satisfaction with life, and mental well-being scales. Descriptive statistics and multiple regression analysis were performed. The data were rechecked and analyzed with the R programming language

**Results:** The prevalence estimates of mental well-being and satisfaction with life were 27% and 13%, respectively. In a total of 660 participants, 58.2% of them were male and the rest of them were female (41.8%). Among the participants, 22.5% suffer the worst conditions regarding their financial conditions, and 16.5% badly seek a job for livelihood.

**Conclusion:** The present findings revealed that the COVID-19 pandemic and longtime educational institution closure significantly affect the students mental health. Students’ mental well-being was in vulnerable conditions and their satisfaction with life was extremely poor. A comprehensive student psychological support service should be expanded to help students’ mental health.

## 1. Introduction

The global health catastrophe caused by the COVID-19 pandemic poses a serious threat both physically and mentally. SARS-CoV-2, the novel coronavirus that produced the COVID-19 pandemic, has a profound influence on the entire human race, with long-term consequences that are still unknown. The following COVID-19 pandemic, and the people’s psychological capacity to cope with a prolonged crisis is challenging. Besides inflicting mortality worldwide, this pandemic also creates psychological pressure on vulnerable populations, especially students, who are already suffering from various stressors (depression, loneliness, anxiety, and stress) (Tasnim et al., 2020).

Many countries imposed travel bans, social gathering restrictions, and educational institution’s closures after the virus spread fast and widely in a relatively short period of time (Rabi et al., 2020). For millennia, quarantine has been used as a preventive tool for the large infectious epidemics worldwide, and it has been proven to be successful in reducing the spread of contagious illnesses like cholera and plague in the past (Brooks et al., 2020). All educational institutes, entertainment, and other public facilities, including restaurants, movie theaters, gyms, shopping malls, and places of worship, were closed a few days later, prohibiting the free movement of people across borders without special authorization. Students’ lives have changed considerably in a short amount of time as they have been ordered to leave school and adjust to new living situations. Students have to leave their campus immediately after the declaration of country-wide lockdown and go back to their respective areas again for an uncertain period of time. It could lead to a high rate of depression, anxiety, stress, and other mental health problems among students (Roy et al., 2020), which may have life-long consequences.

Moreover, for prospective students who come from other countries and join an educational institute, making new social relationships can be extremely difficult, and this can lead to feelings of loneliness or disconnection. Loneliness has been linked to increased stress, anxiety, depression, and other mental health burdens among students, impacting mental well-being (Richardson et al., 2017) and satisfaction with life. The rapid transmission of novel coronavirus COVID-19 has changed this landscape dramatically, from instructional delivery to campus closures, contributing to an already complicated combination of factors affecting student well-being (Burns et al., 2020) and life satisfaction.

In addition, after a few days of university closure, the students were asked to adapt to online learning platforms. Students’ stress levels are likely to have grown due to the shift to online learning, particularly in courses that were not originally planned for online delivery (Kecojevic et al., 2020). Courses which need physical presence, physical labs, internship, and artistic performances, have distinct disadvantages when delivered online (Sahu, 2020). Besides, it’s assumed that some students have trouble accessing to computers and internet at home, specially students from low-socio economic backgrounds and living in rural areas (Hernandez, 2019; Sahu, 2020).

Moreover, concerns about individual health, the health of family members, and, constraints on movement and outdoor recreation,, self-isolation, quarantine, disrupted ordinary daily activities, affect overall mental well-being, financial status, particularly those students who support themselves by working,and overall life satisfaction of this group of people (Di Renzo et al., 2020; Đogaš et al., 2020; Kriaucioniene et al., 2020). These new circumstances, together with the general sense of ambiguity, resulted a widespread distress with a negative influence on the psychological health, as seen by an increase in reported sadness and anxiety among the people (Brooks et al., 2020). As a result of these mental health problems, unhealthy habits may emerge as coping techniques (Pfefferbaum & North, 2020). Students’ academic progress and social connections can be greatly harmed by these mental health issues, limiting their future professional and personal potentiatlity.

Furthermore, several prior studies among Bangladeshi students have repeatedly noted asignificant mental repercussions specially among university students (Anjum et al., 2019; Cao et al., 2020; Hossain et al., 2020; Konstantinovs & Lapa, 2021). As a result of the suspension of educational activities, together with the disruption of regularity and restricted human interactions, 83% believe that their already existing mental disorders have been aggravated (Konstantinovs & Lapa, 2021).

However, in low- and middle-income countries (LMICs), such as Bangladesh, where little resources are available to combat both COVID-19, the mental health concerns is more prevalent as a result of this outbreak. Additionally, there is a dearth of research evaluating the mental well-being and satisfaction with life of students in Bangladesh during the pandemic. To bridge that gap, this study sought to investigate the prevalence and determinants of mental well-being and satisfaction of life among the students.

## 2. Methods

### 2.1 Participants, Study procedure, and Measures

A cross-sectional study was employed to assess mental well-being and satisfaction with life among the students. The purposive sampling technique is also utilized in the present study. The survey was conducted from February to April 2021. The inclusion criteria for the study were i) being a university student, ii) >18 years of age, and iii) has the ability to read and speak Bangla.

A semi-structured self-reported e-survey questionnaire (in Bangla) was developed and an easily accessible Google survey form was created and publicly circulated on multiple social media platforms (Facebook, WhatsApp, etc.). Sufficient number of research assistants were recruited to get a high response rate in the survey.

All participants provided informed consent after the purpose and objectives of the study were thoroughly explained to them. Participation in this survey was voluntary and anonymous.

Before going on to the next phase, a pilot test was undertaken with 50 participants from the same population (target group) to ensure that the questionnaire was acceptable and transparent. Initially, there were 855 responses, and after removing incomplete responses, there were 660 responses for final analysis. The e-questionnaire consisted of three sections: socio-demographic, mental well-being, and satisfaction with life amidst COVID-19 during the last month.

### 2.2 Socio-demographic measures

Socio-demographic characteristics were collected by asking age, sex (male/female), relationship status (single/ engaged/ married), family type (nuclear/joint family), residence (urban/rural), monthly family income (<15,000 Bangladeshi Taka (BDT), 15,000–30,000 BDT, and >30,000 BDT) (Islam et al., 2020). Crisis during COVID-19 were measured by asking the following questions: “are you currently searching for a job?” (trying/ moderately trying/ crying need/ not trying), “your financial conditions during COVID-19” (good/ better/ best/ worst), “your relationships with loved ones (good/ better/ best/ poor).

### 2.3 Satisfaction with life scale (SWLS)

The satisfaction with life scale is the most widely used scale to measure life satisfaction (Yun et al., 2019). The scale consists of five items, with which respondents indicate their level of agreement or disagreement on a seven-point Likert scale (from 7 = Strongly agree to 1 = Strongly disagree). Total scores ranged from 5 to 35, with the lowest scores indicating Extremely dissatisfied (scores between 5 and 9), scores between 10 and 14 indicating dissatisfied, 19-19 indicating Slightly dissatisfied, 20 indicating Neutral, 21-25 indicating slightly satisfied, 26-30 indicating Satisfied, and 31-35 indicating Extremely satisfied. The SWLS has demonstrated satisfactory psychometric properties, a significant degree of internal consistency (Cronbach’s varying from 79 to 89 in various studies), and a greater level of chronological reliability (Rogowska et al., 2021), (Aishvarya et al., 2014). In the present study, the SWLS found to be have very good reliability (Cronbach’s alpha =0.88)

### 2.4 Warwick-Edinburg mental well-being scale (WEMWBS)

WEMWBS is a mental well-being metric that focuses solely on positive aspects of mental health. It holds promise as a tool for monitoring mental well-being at the population level because it is a short and psychometrically robust scale with no ceiling effects in a population sample (Tennant et al., 2007). An expert panel developed it based on current academic literature, qualitative research with focus groups, and psychometric testing of an existing scale. The scale consists of fourteen items, with a five-point Likert scale (from 1 = None of the time to 5 = All of the time). The Likert scale assigns a score of 1 to 5 to each item, with a minimum score of 14 and a maximum score of 70. All items received a positive score. The WEMWBS overall score is calculated by adding the scores for each item with equal weights. As a result, a higher WEMWBS score indicates a higher level of mental well-being. In the present study, the WEMWBS showed content reliability (Cronbach’s alpha =0.89)

### 2.5 Sampling procedure

The sample size was calculated using the following equation:

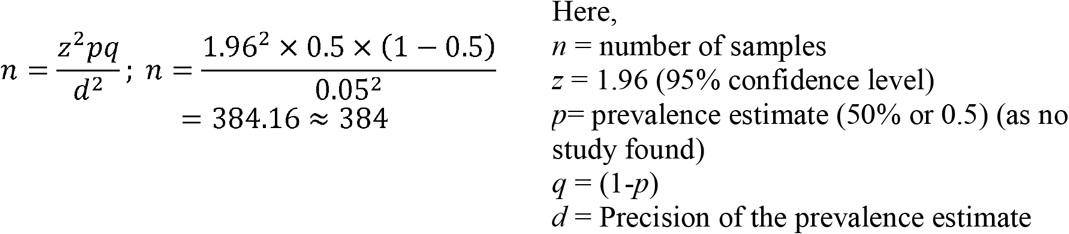

The calculated sampling size was 384. There are limited studies to base this on however, p=0.5 was initially selected. Our sample size exceeds this by a substantial proportion. Out of 855 received responses, 660 responses were analyzed after removing incomplete or ineligible data.

### 2.6 Statistical analysis

Descriptive and inferential statistics (such as frequencies, percentages, means, and standard deviations) were computed. Inferential statistics included using t-tests or one-way Analysis of Variance (ANOVA) to determine mean differences in mental well-being scores in relation to background variables. Skewness, Kurtosis, and Pearson correlations were calculated between all measures pertaining to mental well-being and life satisfaction. The Cronbach’s alpha for SWLS and WEMWBS were 0.88 and 0.89 respectively. And then a well fitted regression model was used to determine relationship between SWLS and WEMWBS with demographic variables. All analyses were carried out with a p-value of < 0.05 using the R programming language.

### 2.7 Ethics

The present study was carried out in accordance with Institutional Research Ethics and the Helsinki Declaration. This study was approved by the respective Ethical Review Committee[Ref: UAMC/ERC/27/2021].. The study’s objectives were explicitly defined in the first part of the questionnaire, along with i) the current research processes, ii) data confidentiality and privacy, and iii) the right to withhold data from the study at any time.

## 3. Results

### 3.1 Descriptive analysis

In this study, we use age, sex, relationship status, family type, residence, monthly family income, currently searching for a job, financial situation during COVID-19, and relationships with loved ones as personal variables. Age 18-23 years (75.9%), male (69.5%), marital status in a relationship (72.1%), joint family (71.7%), rural residence (69.5%), monthly family income <15,000 BDT (69.6%), searching job as a crying need (73.6%), bad financial situation during COVID-19 (81.2%), bad relationships with loved ones (82.5%) are good satisfaction with their life that means their score more than 13 in SWL scale**[Table:2]**. Male those age 18-23 years (43%), Unmarried (38.6%), Joint family (43%), rural residence (46%), family income 5,000-30000 BDT (39%), currently searching for a job as yes trying (42.8%), better financial condition during covid-19 (32.3%), best relationships with loved ones (34.1%) are good satisfied with their life **[Table:1]**. Female those one age 30-35 years(42.9%), unmarried (25.7%), nuclear family (28.1%), urban residence (30%), monthly family income >30,000 BDT (34%), currently searching for a job as moderately trying (35.7%), best financial situation during covid-19 (36.6%), poor relationships with loved ones (37.5%) are not satisfied with their life **[Table:1]**. Age 18-23 years (51.6%), female (53%), relationship status in a relationship (78.2%), joint family (62.8%), rural residence (81.9%), monthly family income <15,000 BDT (50.1%), currently searching for a job as moderately trying (87.9%), best financial situation during covid-19 (68.8%), best relationships with loved ones (56.7%) are good wellbeing during last month **[Table:3]**.

**Table-1:** General characteristics of personal variables.

| Variables |  | Prevalence (%) | Prevalence of Satisfaction With Life (score≥13) |  | Prevalence of mental well-being (score≥27) |  |
| --- | --- | --- | --- | --- | --- | --- |
|  |  |  | Male (58.2%) | Female (41.8%) | Male (58.2%) | Female (41.8%) |
| Age | 18-23 | 419 (67.1%) | 180 (43%) | 107 (26%) | 131 (31%) | 104 (25%) |
|  | 24-29 | 220 (30.1%) | 83 (37.7%) | 61 (27.7%) | 54 (24.5%) | 50 (22.7%) |
|  | 30-35 | 21 (2.9%) | 6 (28.6%) | 9 (42.9%) | 4 (1.9%) | 6 (28.6%) |
| Relationship status | Unmarried | 440 (60.1%) | 170 (38.6%) | 113 (25.7%) | 112 (25.5%) | 77 (1.8%) |
|  | In a Relationship | 140 (19.1%) | 62 (4.4%) | 54 (3.9%) | 42 (3%) | 37 (26%) |
|  | Married | 149 (20.4%) | 35 (23%) | 39 (25%) | 33 (22%) | 46 (3%) |
| Family type | Joint | 180 (24.6%) | 78 (43%) | 51 (28%) | 61 (34%) | 52 (29%) |
|  | Nuclear | 544 (74.3%) | 190 (35%) | 155 (28.1%) | 127 (23%) | 107 (2%) |
| Residence | Urban | 463 (63.4%) | 146 (32%) | 145 (30%) | 103 (22%) | 108 (23%) |
|  | Rural | 259 (35.4%) | 118 (46%) | 61 (24%) | 82 (32%) | 52 (2%) |
| Monthly family income | <15,000 BDT | 168 (23%) | 83 (5%) | 33 (2%) | 61 (36%) | 23 (14%) |
|  | 15,000–30,000 | 276 (37.7%) | 107 (39%) | 77 (28%) | 68 (25%) | 56 (2%) |
|  | >30,000 BDT | 281 (38.4%) | 789 (28%) | 96 (34%) | 60 (21%) | 80 (28%) |
| Currently searching for a job? | trying | 250 (34.2%) | 107 (42.8%) | 48 (19.2%) | 62 (24.8%) | 30 (12%) |
|  | moderately trying | 182 (24.9%) | 51 (28.02%) | 65 (35.7%) | 47 (25.8%) | 55 (30.2%) |
|  | crying need | 121 (16.5%) | 51 (42.1%) | 38 (31.4%) | 44 (33.9%) | 36 (29.8%) |
|  | not trying | 176 (24%) | 59 (33.5%) | 55 (31.2%) | 36 (20.5%) | 39 (22.2%) |
| Financial situation during COVID-19 | good | 153 (20.9%) | 55 (35.9%) | 26 (17%) | 29 (19%) | 13 (8.5%) |
|  | better | 316 (43.2%) | 102 (32.3%) | 91 (28.8%) | 80 (25.3%) | 74 (23.4%) |
|  | best | 93 (12.7%) | 35 (37.6%) | 34 (36.6%) | 33 (35.5%) | 31 (33.33%) |
|  | worst | 165 (22.5%) | 77 (46.7%) | 56 (33.9%) | 47 (28.5%) | 42 (25.5%) |
| Relationships with loved ones | good | 244 (33.3%) | 86 (35.2%) | 57 (23.4%) | 47 (19.3%) | 34 (13.9%) |
|  | better | 186 (25.4%) | 78 (41.9%) | 48 (25.8%) | 52 (28%) | 43 (23.1%) |
|  | best | 255 (34.8%) | 87 (34.1%) | 84 (32.9%) | 73 (28.6%) | 68 (26.7%) |
|  | worst | 40 (5.5%) | 18 (45%) | 15 (37.5%) | 16 (40%) | 14 (35%) |

**Table-2:** Association between personal variable and satisfaction with life (SWL).

| Variables | | Prevalence of satisfaction with life (score≥13) | OR (95% of CI) | P value |
| --- | --- | --- | --- | --- |
| Age | 18-23 | 318 (75.9%) | .77 (.27-2.19) | .624 |
|  | 24-29 | 144 (65.5%) | .88 (.30-2.55) | <.001 |
|  | 30-35 | 15 (71.4%) | 1 |  |
| Sex | Male | 269 (69.5%) | .72 (.49-1.04) | .040 |
|  | Female | 207 (68.5%) | 1 |  |
| Relationship status | Unmarried | 284 (64.5%) | .99 (.62-1.58) | .051 |
|  | In a Relationship | 101 (72.1%) | .66 (.54-1.71) | .899 |
|  | Married | 89 (59.7%) | 1 |  |
| Family type | Joint | 129 (71.7%) | 1 |  |
|  | Nuclear | 346 (63.6%) | .82 (.54-1.24) | .355 |
| Residence | Urban | 191 (42.3%) | .75 (.51-1.11) | .158 |
|  | Rural | 180 (69.5%) | 1 |  |
| Monthly family income | <15,000 BDT | 117 (69.6%) | 1.15 (.66-1.96) | .590 |
|  | 15,000–30,000 | 184 (66.67%) | 1.30 (.86-1.95) | <.001 |
|  | >30,000 BDT | 174 (61.9%) | 1 |  |
| Currently searching for a job? | trying | 156 (62.4%) | .91 (.57-1.46) | .715 |
|  | moderately trying | 116 (63.7%) | .72 (.43-1.20) | .216 |
|  | crying need | 89 (73.6%) | 1.08 (.58-2.01) | .792 |
|  | not trying | 114 (64.8%) | 1 |  |
| Financial situation during COVID-19 | good | 81 (52.9%) | .33 (.18-.59) | <.001 |
|  | better | 193 (61.1%) | .38 (.22-.63) | <.001 |
|  | best | 69 (74.2%) | .79 (.37-1.66) | .538 |
|  | worst | 134 (81.2%) | 1 |  |
| Relationships with loved ones | good | 143 (58.6%) | .24 (0.08-.74) | .013 |
|  | better | 127 (68.3%) | .31 (.10-.95) | <.001 |
|  | best | 171 (67.1%) | .29 (0.09-.89) | <.001 |
|  | worst | 33 (82.5%) | 1 |  |

**Table-3:** Association between personal variable and mental well-being.

| Variables | | Prevalence mental well-being (score≥27) | OR (95% of CI) | P value |
| --- | --- | --- | --- | --- |
| Age | 18-23 | 216 (51.6%) | 1.35 (.48-3.81) | .569 |
|  | 24-29 | 104 (47.3%) | 1.26 (.43-3.36) | **<.001** |
|  | 30-35 | 10 (47.6%) | Reference |  |
| Sex | Male | 189 (44.4%) | .69 (.45-.95) | **<.001** |
|  | Female | 160 (53%) | Reference |  |
| Relationship status | Unmarried | 190 (67%) | .87 (.56-1.36) | **<.001** |
|  | In a Relationship | 79 (78.2%) | Reference |  |
|  | Married | 79 (53.02%) | 1.31 (.74-2.33) | .348 |
| Family type | Joint | 113 (62.8%) | Reference |  |
|  | Nuclear | 235 (43.2%) | .46 (.30-.67) | **<.001** |
| Residence | Urban | 134 (28.9%) | .77 (.52-1.14) | .162 |
|  | Rural | 212 (81.9%) | Reference |  |
| Monthly family income | <15,000 BDT | 85 (50.1%) | .77 (.88-1.63) | .935 |
|  | 15,000–30,000 | 124 (44.9%) | .81 (.54-1.22) | **<.001** |
|  | >30,000 BDT | 140 (49.8%) | Reference |  |
| Currently searching for a job? | trying | 92 (58.33%) | .75 (.45-1.15) | .172 |
|  | moderately trying | 102 (87.9%) | 1.25 (.76-2.05) | .366 |
|  | crying need | 81 (66.9%) | 1.51 (.88-.72) | **<.001** |
|  | not trying | 75 (42.6%) | Reference |  |
| Financial situation during COVID-19 | good | 42 (27.5%) | .17 (.27-.81) | **<.001** |
|  | better | 155 (49.1%) | .70 (.45-1.11) | **<.001** |
|  | best | 64 (68.8%) | 1.49 (.76-2.29) | .122 |
|  | worst | 89 (53.9%) | Reference |  |
| Relationships with loved ones | good | 81 (33.2%) | .23 (0.09-.58) | .002 |
|  | better | 95 (51.1%) | .35 (.14-.88) | **<.001** |
|  | best | 142 (56.7%) | .38 (.15-.94) | **<.001** |
|  | poor | 30 (75%) | Reference |  |

### 3.2 Association among satisfaction with life, mental well-being during last month, and personal variables

Those with better financial situation during COVID-19 [odd ratio: 38%, CI (.22-.63), p value < 0.05] and best relationships with loved ones [OR: 29; 95%CI (0.09-.89), p value < 0.001] are good satisfied with their life comparing to others **[Table: 2]**. Age [OR: 88; 95% CI (.30-2.55), p value < 0.001], Male [OR: 69; 95% CI (.45-.95), p value < 0.001], nuclear family type [OR: 46; 95% CI (.30-.67), p value < 0.001], good financial situation during COVID-19 [OR:17; 95% CI (.27-.81), p value <0001], good relationships with loved ones [odd ratio .23, 95% of CI (0.09-.58), p value < 0.001] are good mental well-being during last month comparing to others **[Table:3]**. Overall, Those aged 18-23 years, relationship status in a relationship, joint family, rural residence, monthly family income <15,000 BDT, poor relationships with loved ones are simultaneously soundly satisfied with life and mental well-being during last month are good **[Table:1]**.

## 4. Discussion

The global pandemic has significantly transformed the student welfare sector partly because of extensive pragmatic reforms to limit the spread of COVID-19. Across the board, education has seen significant changes in how education is delivered. The shift to online learning has happened quickly, posing a variety of new issues for both faculty and students. The required transformation in educational institutions has had a significant (mostly unfavorable) influence on students’ overall learning experience and psychological well-being. Therefore, a study regarding their mental well-being and life satisfaction become necessary to address the issues. However, the objective of this study was to determine the mental well-being and satisfaction with life of students during COVID-19 outbreak in Bangladesh. To the best of author’s knowledge this is the first study reporting students mental well-being and satisfaction with during the COVID-19 pandemic in Bangladesh.

According to the present findings, students reported low mental well-being and poor life satisfaction. In addition, the present study reported male students, early adults, middle socio-economic status, and job seeker students are significantly more vulnerable to poor mental health and life satisfaction. As per the study, students’ mental health is jeopardized during the pandemic. This study reports the mental well-being and satisfaction with life of students in Bangladesh while the education sector was substantially disturbed by COVID-19. However, the present study revealed that only 13% of students are satisfied with their lives, and the prevalence of mental well-being is 27%.

Despite differences in survey populations, methods, and cultures, the current findings are comparable to previous research on satisfaction with life and mental well-being and related factors in students and other populations.

However, a recent study conducted among nine countries university students found that higher satisfaction with life (60.54%). However, Columbia reported the highest satisfactory (81.94%) students and on the other hand Turkey reported the lowest prevalence of life satisfaction (28.06%) of students (Rogowska et al., 2021). Moreover, a recent study in Germany revealed that 72.2% students are suffering from serious impairment of mental well-being (Brähler et al., 2007), and another study revealed that 75.8% have serious indication of mental disorder (Holm-Hadulla et al., 2021).

However, the early adults (24 to 29 years) had significantly poor life satisfaction in this study. A prior study indicated that as people grew older, they became less satisfied with their lives (Chen, 2001). While determining good well-being, this study showed that the prevalence of good mental well-being found significantly associated with middle age people. In line with the present study another study among students reported that increased age was associated with enhanced psychological well-being (Franzen et al., 2021). A prior study among English and Scottish adolescent students showed no correlation between mental health and age (Clarke et al., 2011). In a survey of students of health disciplines, increased age was found to have a positive association with psychological well-being (Franzen et al., 2021).

However, in the present study, it has been found that male students have a significant association with good mental well-being. In contrast to a study in Denmark among general practitioners, which reported that male were more likely than female to experience poor mental well-being (Nørøxe et al., 2018). Another study of health science university students found no significant associations between and within age groups when it came to their mental well-being (Alshehri, 2021). A similar study among medical students indicated that women’s burnout rates are higher than men (Wimberly et al., 2020). A further study among public health students showed that the psychological discomfort among female was larger than in the general population at the same age (Bíró et al., 2011), indicating the male students as more mentally sound. However, a previous study indicated that males are less likely than females to seek care for mental health issues, resulting in greater mental health burden (Sagar-Ouriaghli et al., 2020). Male students, in comparison to female students, have more negative attitudes toward psychiatric services and are less likely to seek treatment (Sagar-Ouriaghli et al., 2020). There is a dearth of evidence-based research to address this issue, despite significant interest. To completely comprehend the psychological impact of COVID-19 on male students, more research is needed.

Moreover, satisfaction with life found significantly associated with monthly family income in this study. According to a previous study, relative income was found to be more important for life satisfaction (Wolbring et al., 2013). Furthermore, students with a better financial standing were found to have higher levels of life satisfaction. Similarities have been found with a study conducted among college students which suggests that favorable financial behaviors contribute to financial satisfaction, which in turn adds to life satisfaction (Xiao et al., 2009). Other research involving university students indicated that when the financial stress of paying tuition is removed, students’ life satisfaction improves dramatically (Slavinski et al., 2021). Consequently, students with higher socio-economic status are more satisfied with their life (Chow, 2005).

Additionally, millions of individuals have lost their jobs as a result of global economic instability following COVID-19 (Crayne, 2020). As a result, these same people will be dealing with the pain of job loss in the present, as well as the stress of job hunting in the future. In addition, the satisfactory relationship with loved ones has also been found to be significantly associated with life satisfaction compared to others. However, the present study is in line with a study among University Students in a Canadian Prairie City (Chow, 2005). A study in Barbados also showed similar findings (Alleyne et al., 2010). According to another study on Family Functioning and Life Satisfaction, people who rate their family functioning as cohesive, adaptable, communicative, and fulfilling are more likely to process their own emotions and have better life satisfaction (Szcześniak & Tułecka, 2020). As a representation of the quality of life, the home is more than just a house. It can offer a variety of advantages for a person’s bodily and psychological well-being and satisfaction with life. So the relationship with family members, especially during the pandemic situation, as well as the relationship with friends and relatives, also plays an important role in mental health.

Nonetheless, each individual’s mental well-being is critical to their performance and productivity. But various stressors and environmental variables can contribute to an imbalance. Students frequently experience mental health issues as a result of recent changes in the education system, online classes, and financial strain. Unmarried people have been found to have good mental well-being in this current study. In contrast to a previous study, which revealed that married people have the highest level of subjective well-being (Dush & Amato, 2005). Longitudinal research, on the other hand, consistently shows that marriage promotes mental well-being (Dush & Amato, 2005; Williams, 2003). As COVID-19 hits different people in different ways, the unmarried participants were found to have good mental well-being as they didn’t need to think regarding their family as they don’t have spouses and childrens.

However, industrialization and globalization have accelerated social transformation, which has affected not just people’s professional life but also their personal lives, particularly in developing countries. As a result, extended families are turning into nuclear families. In the course of time, families were shown to be highly linked to the social adaptation and the psychological well-being of an individual (Kellam et al., 1977). In this study, students from nuclear families have been found to have good mental well-being compared to others. This may be due to the nuclear family having a significant impact on the formation of an individual’s personality. An individual is closer to their parents and may have more open and honest discussions with them about their concerns during quarantine, which aids in maintaining their psychological well-being. Nuclear families are also more likely to use emergency rooms and can have the opportunity to give children adequate healthcare (Anderson, 2014). Moreover, the emotional pressure on children with two parents living in a non-violent family is far lower. This maintains students living in the nuclear family in good mental well-being. A study among Joint and Nuclear Family Women revealed that there was a significant difference in marital adjustment and mental health between women from joint and nuclear families (Patel & Zala, 2011).

However, people from middle socio-economic status (15,000 to 30,000 BDT) demonstrated good mental well-being. An earlier studies in Finland found that low income was associated with poor mental health in the men in the study group (Viinamäki et al., 1993). A comparable study among university students found that prolonged financial stress severely impacted students’ psychological well-being by lowering their sense of comprehensibility about their circumstances, as well as their sense of control and self-esteem (Lange & Byrd, 1998). In contrast to many other studies, those who are looking for work as a crying need exhibited good mental well-being in our current study (Bialowolski et al., 2021; Viinamäki et al., 1993).

Consequently, being a student in today’s society is the most challenging task, and the academic system has been more demanding than ever before. Furthermore, throughout the pandemic, it has become increasingly complex, with increased competitiveness, resulting in increased levels of stress among students. The spread of COVID-19 caused a severe change to the emotional, physical, mental, social, and financial conditions of billions of persons. The COVID-19 pandemic could have a major effect on the mental well-being of many people, especially students. Another prior study at the time of COVID-19 has found that the threat of COVID-19 has a negative impact on subjective mental well being (Meo et al., 2020; Paredes et al., 2021), serial mediation studies revealed that during COVID-19, intolerance, and uncertainty had a large direct effect on mental well-being and satisfaction with life (Satici et al., 2020). According to a recent study, the lockdown, social distancing, and self-isolation requirements are stressful and harmful to many people, causing health, mental well-being, and satisfaction with life problems among students (Nurunnabi et al., 2020).

However, the present study reported that students who have a good relationship with their loved one are found to be good mental well-being and sufficient life satisfaction than others which is in line with a prior study (Viejo et al., 2015). A previous study demonstrated that participants who reported poor family support had alow mental well-being (Cano et al., 2003). In addition, a further longitudinal study is necessary to establish a strong link between relationship with loved ones and mental well-being and life satisfaction.

### Limitations

There are several limitations to the study that should be considered. The main limitation of this study is that participation in the study required access to a smartphone/computer, implying that respondents from the lower socioeconomic subgroup could not be included. Second, because this study relied on self-reported data, it was not completely free of recall or reporting bias. Third, because the study was conducted online using a convenience sampling technique, the possibility of selection bias should be considered. Finally, the study’s cross-sectional design includes method bias because a causal relationship cannot be accurately elucidated in this design. Future qualitative and longitudinal studies will be required to determine the true scenario in the context of the COVID-19 pandemic.

## 5. Conclusions

The current study found that the closing of educational institutions generated significant disturbance in students’ mental health. The COVID-19 pandemic had a significant impact on students’ mental well-being and life satisfaction, and precautions were put in place to prevent its spread. Student’s psychological services in hard-to-reach places should be expanded by the government and other policymakers. The evidence so far in respect to pre-existing models of well-being shows that the pandemic’s psychological influence will be extensive. While students will be living with uncertainty about their studies for an undetermined amount of time, researchers should move rapidly to assess student well-being and life satisfaction in these unprecedented times and beyond.

## Data Availability

All data produced in the present study are available upon reasonable request to the authors.

## Acknowledgements

We thank all the participants who took part in the study. We also acknowledge the efforts of all research assistants that helped in data collection for the study.

## Funding

The authors did not receive any financial support from any public/private organizations or other funding agencies for this study.

## Data Availability

The data that support the fndings of the current study are available from the corresponding author upon reasonable request.

## Conflict of interest

All authors declare that they have no potential conflict of interest in the dissemination of the study’s findings.

## Notes

### Competing Interest Statement

The authors have declared no competing interest.

### Funding Statement

This study did not receive any funding

### Author Declarations

This study was approved by the respective Ethical Review Committee of Uttara Adhunik Medical College[Ref: UAMC/ERC/27/2021].

